# Exploration of the molecular origins of sex-specific and temporal comorbidity patterns in dementia: insights from the Austrian claims data

**DOI:** 10.64898/2026.07.14.26357961

**Authors:** Vladimir Kovačević, Bojana Bašaragin, Jovana Kovačević, Anđelka Zečević, Salvo Danilo Lombardo, Elma Dervic

**Author notes:** (Corresponding author: Elma Dervić.). contributed equally as last authors (Salvo Danilo Lombardo, Elma Dervic).

## Abstract

Dementia is a progressive condition that impairs cognitive processes such as memory, decision-making, and the ability to manage daily activities. Recent estimates suggest that more than half of all dementia cases could be preventable by addressing their risk factors, including disease comorbidities such as diabetes and vision loss. Yet, we lack a comprehensive molecular map of dementia comorbidities. In this work, we analyzed Austrian nationwide hospital claims data, comprising 13 million hospital stays from 2015 to 2019, to systematically assess dementia-related risk across disease comorbidity patterns, covering both their molecular relationships and their epidemiological overrepresentation. We identified disease trajectories occurring before and at the time of dementia diagnosis, revealing both sex-specific and shared comorbidity patterns. Overall, we identified 51 potential risk factors, with a prominent contribution from endocrine and metabolic disorders. While Parkinson’s disease emerged as a strong molecularly related driver of dementia, we also identified emerging and previously under-chracterized risk factors, including vitamin D deficiency. This integrative framework provides a comprehensive view of dementia-associated disease networks and identifies novel, potentially modifiable risk factors. These results offer new opportunities for targeted prevention strategies and advance our understanding of the complex interplay between comorbidities and dementia development.

## I. Introduction

Dementia is a major global health challenge, characterized by progressive decline in cognitive functions such as memory, reasoning, and the ability to perform daily activities [1]. Its impact extends beyond individuals to families, healthcare systems, and societies, with substantial and growing economic and social costs [1], [2]. In 2019, the worldwide costs of dementia were estimated at over 1 trillion USD, a figure projected to rise sharply with demographic aging [2]. The global prevalence of dementia is increasing rapidly. According to a systematic review by Prince et al. (2013), dementia affects approximately 5–7% of adults aged 60 and over, with case numbers expected to nearly double every two decades, rising from 35.6 million in 2010 to an estimated 115.4 million by 2050 [3]. More recent estimates from the Global Burden of Disease Study 2019 indicate that 57.4 million people were living with dementia in 2019, with numbers expected to rise to 152.8 million by 2050 [4]. These projections highlight substantial gender differences, with women more affected than men (female-to-male ratio of 1.67) [4]. Although these studies should alert the scientific and medical communities, many of these cases may be preventable. The 2020 Lancet Commission concluded that about 40% of dementia cases could be preventable or delayable by targeting 12 modifiable risk factors across the life course [5], and recent estimates suggest that this number could be extended to more than half of all dementia cases [6].

Despite being promising, prevention strategies require a deeper understanding of dementia and its associated comorbidities. Dementia rarely occurs in isolation; it is commonly accompanied by other chronic diseases that complicate diagnosis, treatment, and prognosis. Previous work has documented frequent comorbidities such as cardiovascular, metabolic, and neuropsychiatric disorders [7], [8]. Claims-based analyses, longitudinal progression models, and network approaches have been increasingly employed to capture the multimorbidity patterns in dementia [7]–[10]. Machine learning and artificial intelligence (AI) methods are also emerging as tools to aid in early detection, subtype identification, and risk prediction [11]–[14]. A key limitation of such population-level studies is their ability to identify that conditions co-occur, but lack mechanistic interpretability. There is a need for approaches that augment clinical data with external biological evidence to provide a more insightful interpretation of the complex comorbidity landscape in dementia.

Network science provides a powerful framework to analyze multimorbidity by modeling diseases as interconnected systems. Comorbidity networks capture non-random co-occurrence patterns of diseases, allowing the identification of clusters of conditions that may share underlying risk factors or pathophysiological mechanisms [9]. Integrating such approaches with genetic associations offers an additional layer of insight, linking observed clinical patterns to molecular pathways.

In this study, we leverage Austrian nationwide hospital claims data, comprising 13 million hospital stays from 2015 to 2019 across all hospitals and involving nearly 4 million patients. Within this cohort, 111,501 patients were diagnosed with dementia. This figure represents a subset of the estimated 147,000 people living with dementia in Austria in 2018 (1.66% of the total population), as reported by the Alzheimer Europe 2019 Yearbook [15], since hospital claims capture only those patients who had at least one inpatient admission during the study period [16]. Our cohort is further comparable to a nationwide Austrian prescription-claims analysis that included 70,799 patients treated with anti-dementia drugs and reported a distinct co-medication profile compared with matched controls [17].

We constructed dementia-specific comorbidity networks, and identified sex-specific and shared disease co-occurrences at the time of, and prior to, dementia diagnosis. To further contextualize these findings, we integrated disease–gene associations from publicly available databases and harmonized them with our clinical networks. This investigation allows the large-scale, unbiased, identification of potential risk factors, with potential implications for more targeted, patient-centered healthcare interventions.

## II. Background

### A. Analyzing claims data in dementia research

Administrative claims have long served as a valuable and cost-effective resource for dementia research [18]. Although they cannot replace established clinical tests, claims data-based analyses appear as a useful additional measure for cost-effective risk detection in early dementia screening [19]. Validation studies demonstrate that routinely collected health data can achieve high positive predictive values of 80-90% in appropriate settings [20]. Although sensitivity remains more variable, the authors in [20] report that it is improving over time, following the growing awareness of preclinical cognitive impairment and clinicians’ willingness to formally code the disease.

Methodologically, dementia research using claims data has evolved from traditional statistical approaches toward machine learning methods capable of modeling high-dimensional data, capturing complex relationships between variables and identifying latent structures such as disease subtypes to support personalized risk modeling [11], [14], [19]. These models typically achieve moderate predictive performance [19], while recent approaches exploit temporal patterns in healthcare trajectories using both deep learning and longitudinal modeling techniques, showing promising predictive accuracy and improved characterization of disease progression [10], [21].

In parallel, network-based approaches have emerged as a complementary direction, shifting the focus from prediction to the characterization of multimorbidity patterns. By modeling diseases as interconnected systems, these methods enable the identification of disease trajectories, clustering structures, and potential directional relationships between conditions [9], [22]. Extensions of this approach incorporate statistical analytics to quantify how prior diagnoses influence future disease risk, revealing increased probabilities of dementia following specific conditions such as stroke [23]. Large-scale applications further show that comorbidity patterns in dementia are highly complex and do not form easily separable clusters, highlighting the need for holistic and individualized clinical assessment [7]. Recent work on multilayer comorbidity networks constructed from population-wide data additionally shows that disease interactions can be traced across the lifespan, providing a framework for understanding long-term disease progression and uncovering novel associations [22], [24].

### B. Dementia risk factors

Considerable research has focused on identifying lifestyle, environmental, and clinical risk factors associated with dementia. A comprehensive systematic review and consensus report by the Lancet Commission has demonstrated that cardiovascular health, metabolic disorders, physical inactivity, smoking, and low educational attainment, as well as factors such as sensory impairment, social isolation, and environmental exposures (e.g., air pollution), significantly increase the risk of developing dementia [5].

Meta-analyses have quantified these associations, indicating that diabetes is associated with substantially increased dementia risk (1.5-to 2-fold depending on subtype) [25], whereas midlife hypertension shows more modest but still significant associations, particularly with Alzheimer’s disease [25], [26]. Hearing loss in midlife accounts for approximately 8% of dementia cases, making it the largest potentially modifiable risk factor at that stage of life [5]. Other sensory impairments have also been implicated, with the combination of dual sensory impairment (both hearing and vision loss) linked to a 52% increased risk of all-cause dementia [27]. Traumatic brain injury (TBI) also has a well-documented relationship with dementia risk, suggesting that repeated or more severe injuries show higher risk ranging from modest (∼1.2-fold) to substantially elevated risks (approaching 2–3-fold in individuals with multiple TBIs), with evidence for Alzheimer’s disease being less consistent [28], [29].

Beyond individual factors traditionally emphasized in epidemiological studies, a growing body of research shows that multimorbidity is significantly associated with dementia risk, especially in cardiometabolic disease clusters [30], [31]. Besides highlighting the cumulative impact of multiple modifiable risk factors, modeling studies suggest that population-level interventions targeting these factors in midlife or earlier could substantially reduce dementia incidence [32]. This finding is supported by cross-sectional evidence that such risk factors are highly prevalent and unequally distributed across sociodemographic groups [33]. Claims data, when linked with sociodemographic and clinical registries, have increasingly been used to operationalize these risk factors in large populations, allowing for risk stratification and early identification of individuals at elevated dementia risk [11], [19].

### C. Dementia in gene–disease associations

Genetic and molecular research has complemented epidemiological studies by shedding light on the biological causes of dementia and its comorbidities. Genome-wide association studies (GWAS) have identified numerous loci linked to Alzheimer’s disease, vascular dementia, and related cognitive disorders, highlighting pathways involved in amyloid processing, tau pathology, lipid metabolism, and neuroinflammation [34]–[36]. Integrative approaches combining GWAS results with disease networks have revealed that genes implicated in dementia often overlap with those associated with cardiovascular, metabolic, and psychiatric conditions, providing mechanistic insight into observed comorbidity patterns [37], [38].

Furthermore, publicly available databases such as Dis-GeNET and the GWAS Catalog, both of which include extensive Alzheimer’s disease and dementia-related data, enable the systematic integration of gene–disease associations with clinical data [39], [40]. Such integrations offer opportunities to infer potential molecular drivers of disease clusters observed in population-level claims analyses and facilitate the translation of these findings into biologically informed hypotheses for targeted interventions and precision medicine approaches.

Unlike databases that primarily curate published associations, Open Targets [41] combines multiple lines of evidence, including genetic associations from GWAS, text-mined literature, somatic mutations, and RNA expression into a single overall association score which reflects the confidence in each gene–disease link. This evidence-based scoring system enables researchers to prioritize associations with greater confidence and to compare the relative strength of evidence across different gene–disease pairs [42], providing a basis for downstream integration with comorbidity patterns derived from clinical claims data.

## III. METHODS

### A. In-hospital Data

The dataset covers all hospital admissions in Austria from January 2015 to December 2019, a total of 13 million cases from nearly 4 million individuals (N = 3,999,832) [43]. Health insurance is compulsory in Austria, and the share of people without coverage was estimated at about 0.3% in 2015 [44]. The dataset includes sex, age in five-year intervals, and detailed records of each hospital stay: patient ID, primary and secondary diagnoses, admission and discharge dates, discharge type, hospital department, residence and hospital regions, and nationality. Diagnoses are coded using level-4 ICD-10, a global standard maintained by the WHO, enabling consistent classification across 12,040 unique diagnoses. In this study, we use ICD-10 3-digit codes from A to N, codes of diseases, in total 1080 codes. Furthermore, primary (main reason for hospital admission) and secondary diagnoses (all present conditions during hospital admission) are studied as equally important. The dementia cohort included patients with at least one hospital admission (2015–2019) in which F00, F01, F02, F03, or G30 appeared as a primary or secondary diagnosis.

### B. Comorbidity Network Construction

Comorbidity or disease-disease networks capture how different diseases are interconnected, often revealing patterns not visible when conditions are studied in isolation. Built from large-scale longitudinal health datasets, these networks rely on statistical tools and concepts from network science to uncover meaningful (often temporal) disease correlations. To quantify how strongly two diseases are linked, we use Odds Ratios (OR)-a simple measure for network construction [24]. To account for confounders like age, sex, and time, we apply the Cochran-Mantel-Haenszel (CMH) method, which stratifies data and computes a weighted average of ORs, offering more reliable insights into true disease relationships [45]. We constructed contingency tables for each disease pair for each stratum (10-year age group for every two years for females and males). An edge between two diagnoses was retained if the odds ratio exceeded 1.5, the association was statistically significant (p < 0.05), and at least 100 patients had both diagnoses [24]. Only diseases showing a statistically significant association with at least one dementia code (F00,F01,F02,F03,G30) were retained, obtaining a female-specific, dementia-specific network with 173 nodes and 4083 edges, and a male-specific, dementia-specific network with 62 nodes and 1167 edges.

### C. Comorbidity network parameters

Table I provides an overview of the comorbidity network parameters.

### D. Louvain community detection

We used the Louvain algorithm to detect communities within each sex-specific dementia-specific comorbidity network, aiming to identify groups of nodes that are more densely connected to each other than to the rest of the network [46]. The method maximizes modularity, a measure that compares the observed intra-community edge density to that expected under a random null model. It proceeds iteratively in two phases: first, nodes are assigned to communities to maximize modularity locally; second, a new network is constructed where communities become nodes, and the process repeats until convergence. The resolution parameter was set to the one corresponding at the highest modularity, r = 0.9 in females, and r = 1 in males.

### E. Risk factor detection

Risk factor analysis was performed using two complementary temporal frameworks preceding dementia diagnosis: a short-term (2-year) window and a long-term (5-year) window. For the short-term analysis, we quantified disease co-occurrence by computing odds ratios (ORs) within the 2-year period preceding dementia diagnosis. Only diagnoses occurring before the first recorded dementia event were considered, assuring a temporal ordering consistent with potential risk factor identification. For the long-term analysis, we assessed disease incidence patterns spanning age groups by computing z-scores for each ICD-10 diagnosis. Specifically, for each age bracket and sex, we compared the incidence of each disease among individuals who developed dementia to that observed in the overall population. Diseases with a z-score greater than 2 were considered significantly overrepresented within a given age group. To improve robustness, we defined candidate risk factors using both methods. Only diseases consistently over-represented across all age groups (z-score > 2) and showing a short-term association with dementia (OR > 1.5) were kept as potential risk factors. These selected conditions were used to construct a network of dementia risk factors. The network was modeled as a bipartite graph, linking dementia diagnoses (ICD-10 codes: F00, F02, F03, and G30) to their associated risk factors.

### F. Preparing genetic comorbidity signatures and harmonizing with hospital records

We constructed a disease–disease comorbidity matrix using gene–disease association data obtained from the Open Targets Platform [41], [47] (release v25.09). Each entry contains a gene–disease pair along with an overall association score ranging from 0 to 1, summarizing the strength of supporting evidence from multiple sources.

We first filtered out any associations with an overall score of zero, ensuring that only disease–gene pairs with at least minimal supporting evidence were retained. For each disease, we collected its associated genes and their scores into a sparse matrix *X* ∈ ℝ ^*n*×*m*^, where *n* is the number of diseases and m is the number of genes. Each element *X*_*ik*_ represents the association score between disease *i* and gene *k*, or zero if no association is reported. We used Pearson correlation between associated genes as similarity measure between diseases. This scoring method ensures that not only the overlapping gene associations contribute, but also the scores. Self-comparisons (*i* = *j*) are excluded by setting the diagonal to zero. This comorbidity matrix can be used for further network analysis or to identify clusters of diseases with shared genetic underpinnings.

To integrate literature-independent genetic associations with standardized clinical diagnostic codes, we harmonized a disease–disease association matrix from Open Targets with ICD-10 disease nomenclature. PubMedBert [48], a pre-trained biomedical sentence embedding model, was used to semantically map disease names in the Open Targets dataset to the most similar ICD-10 descriptions. For each disease, we computed cosine similarity between its embedding and those of all ICD-10 terms, accepting the best match if the similarity exceeded a threshold (cosine similarity > 0.8); otherwise, the disease was excluded from downstream analyses. A cosine similarity threshold of 0.8 was adopted, as prior work has shown that the value 0.5 provides the balanced trade-off between precision and recall in embedding-based semantic retrieval [49] we wanted to make sure that precision is boosted maximally and avoid false connections.

Following the mapping, the association matrix was filtered to retain only diseases with confident ICD-10 mappings, and duplicate ICD-10 labels arising from many-to-one mappings were merged by taking the maximum value across corresponding rows and columns. To enhance alignment with clinical frameworks, we consolidated all dementia subtypes, such as Alzheimer’s disease (G30) and vascular dementia (F01), under a single harmonized representation by computing the element-wise maximum across their network profiles.

## IV. Results

Among 3,999,832 individuals hospitalized in Austria between 2015 and 2019, 111,501 had at least one dementia diagnosis (F00, F01, F02, F03, G30): 69,045 were female and 42,456 were male, Table III. Dementia patients were substantially older than those without dementia (mean age 86.05 vs 51.66 years). Hence, we have (age, sex, and year of dementia diagnosis) matched the two cohorts to get comparable cohorts, Table IV. Dementia patients exhibited higher healthcare utilization, including more hospital stays (mean 4.75 vs 3.89), longer hospitalizations (45.86 vs 27.68 days), and more diagnoses (4.15 vs 3.03). They also showed a high prevalence of cardiometabolic and neuropsychiatric conditions, notably hypertension (I10–I15: 66.02%), atrial fibrillation (I48: 32.44%), diabetes (E10–E14: 24.33%), and depressive disorders (F32–F33: 17.08%). Throughout 2015–2019, dementia was consistently more frequent among females, both in patient numbers and in the proportion of hospital admissions involving dementia, Figure 1. Over time, the female ratio showed a mild decline, while male ratios remained comparatively stable with less year-to-year variation [50]. This might be due to the fact that we have more age bins for females versus males. This sex-specificity is also reported on the different sizes of the dementia-specific co-occurrence networks: females showed a larger number of comorbidities (173 nodes, 4083 edges) compared to males (62 nodes, 1167 edges), Figure 2.

**TABLE I:** Node-level centrality and structural measures computed for each network independently.

| Measure | Definition | Normalisation |
| --- | --- | --- |
| Degree centrality | Number of direct connections per node | Divided by $n - 1$ |
| PageRank centrality | Iterative influence score with damping factor $\alpha = 0.85$ | — |
| Clustering coefficient | Proportion of a node's neighbours that are also connected to each other | — |
| K-core index | Highest-order core in which a node resides | Divided by max k-core |
| Betweenness centrality | Fraction of all shortest paths that pass through a node | Normalised by path count |
| Closeness centrality | Inverse of the mean shortest path length from a node to all others | — |
| Eigenvector centrality | Node influence weighted by the importance of its neighbours | — |

**TABLE II:** Graph-level topological statistics computed for each network. Path-based metrics were computed on the largest connected component (LCC).

| Measure | Definition |
| --- | --- |
| Nodes & edges | Network size and total number of connections |
| Network density | Ratio of observed edges to the maximum possible number of edges |
| Avg. clustering coefficient | Mean of node-level clustering coefficients across the network |
| Average degree | Mean number of connections per node |
| Degree heterogeneity (CV) | Coefficient of variation of degree (standard deviation / mean) |
| Maximum k-core | Depth of the innermost core structure of the network |
| PageRank entropy | Shannon entropy of the PageRank distribution; higher values indicate more uniform node influence |
| Transitivity | Global tendency of nodes to form triangles |
| Degree assortativity | Correlation between degrees of connected node pairs |
| Avg. shortest path (LCC) | Mean shortest path length computed on the largest connected component |
| Diameter (LCC) | Longest shortest path within the largest connected component |
| Connected components | Number of disconnected subgraphs; values $> 1$ indicate network fragmentation |

**TABLE III:** Baseline characteristics for patients with and without a diagnosis of dementia (ICD-10: F00, F01, F02, F03, G30), respectively for the total samples and male and female patients.

|  | Dementia |  |  | Without Dementia |  |  |
| --- | --- | --- | --- | --- | --- | --- |
|  | All | Female | Male | All | Female | Male |
| All patients | 111,501 | 69,045 | 42,456 | 3,888,330 | 2,084,295 | 1,804,035 |
| Age | 86.05+/-7.84 | 87.07+/-7.47 | 84.38+/-8.13 | 51.66+/-23.89 | 51.95+/-23.6 | 51.32+/-24.22 |
| Hospital_stay | 4.75+/-4.2 | 4.52+/-3.95 | 5.13+/-4.55 | 2.78+/-4.14 | 2.73+/-3.97 | 2.84+/-4.32 |
| Hospital_days | 45.86+/-45.61 | 43.89+/-44.96 | 49.06+/-46.46 | 14.84+/-30.31 | 14.47+/-29.76 | 15.27+/-30.94 |
| Number_diagnoses | 4.15+/-2.91 | 3.99+/-2.83 | 4.41+/-3.03 | 2.15+/-1.83 | 2.14+/-1.77 | 2.17+/-1.89 |
| Mortality | 0.24 | 0.21 | 0.29 | 0.04 | 0.03 | 0.05 |
| E66 | 4.19 | 4.38 | 3.89 | 3.83 | 3.71 | 3.97 |
| E78 | 19.63 | 18.82 | 20.96 | 8.06 | 6.74 | 9.58 |
| F10 | 3 | 1.47 | 5.48 | 1.96 | 1.04 | 3.03 |
| F17 | 2.52 | 1.71 | 3.82 | 2.95 | 2.15 | 3.89 |
| F41 | 1.97 | 2.5 | 1.11 | 0.88 | 1.09 | 0.65 |
| G20 | 8.98 | 6.76 | 12.59 | 0.49 | 0.41 | 0.59 |
| I10 | 66.02 | 67.34 | 63.87 | 19.5 | 18.06 | 21.17 |
| I48 | 32.44 | 31.28 | 34.35 | 5.97 | 5.2 | 6.85 |
| S06 | 9.03 | 8.86 | 9.29 | 2.77 | 2.17 | 3.46 |
| F32-F33 | 17.08 | 19.82 | 12.63 | 3.81 | 4.61 | 2.89 |
| G45-G46 | 5.06 | 4.91 | 5.3 | 1.04 | 0.99 | 1.09 |
| I10-I15 | 66.53 | 67.86 | 64.36 | 19.68 | 18.23 | 21.35 |
| E10-E14 | 24.33 | 22.66 | 27.05 | 6.89 | 5.59 | 8.39 |
| I20-I25 | 24.84 | 21.65 | 30.03 | 7.88 | 5.43 | 10.72 |
| H90-H91 | 4.9 | 4.67 | 5.27 | 0.94 | 0.87 | 1.03 |

**TABLE IV:** Baseline characteristics for patients with and without a diagnosis of dementia (ICD-10: F00, F01, F02, F03, G30), matched by age, sex,and year of dementia diagnosis.

|  | Dementia |  |  | Without Dementia |  |  |
| --- | --- | --- | --- | --- | --- | --- |
|  | All | Female | Male | All | Female | Male |
| All patients | 111,501 | 69,045 | 42,456 | 111,501 | 69,045 | 42,456 |
| Age | 86.05+/-7.84 | 87.07+/-7.47 | 84.38+/-8.13 | 85.88+/-7.89 | 86.92+/-7.53 | 84.2+/-8.18 |
| Number of Hospital Stays | 4.75+/-4.2 | 4.52+/-3.95 | 5.13+/-4.55 | 3.89+/-4.8 | 3.72+/-4.71 | 4.15+/-4.93 |
| Number of Hospital Days | 45.86+/-45.61 | 43.89+/-44.96 | 49.06+/-46.46 | 27.68+/-35.09 | 27.61+/-35.41 | 27.79+/-34.57 |
| Number of Diagnoses | 4.15+/-2.91 | 3.99+/-2.83 | 4.41+/-3.03 | 3.03+/-2.35 | 2.95+/-2.29 | 3.17+/-2.44 |
| Mortality | 0.24 | 0.21 | 0.29 | 0.16 | 0.16 | 0.18 |
| E66 | 4.19 | 4.38 | 3.89 | 3.69 | 3.82 | 3.48 |
| E78 | 19.63 | 18.82 | 20.96 | 15.43 | 14.66 | 16.68 |
| F10 | 3 | 1.47 | 5.48 | 0.65 | 0.25 | 1.3 |
| F17 | 2.52 | 1.71 | 3.82 | 1.33 | 0.87 | 2.08 |
| F41 | 1.97 | 2.5 | 1.11 | 0.78 | 1.01 | 0.41 |
| G20 | 8.98 | 6.76 | 12.59 | 2.08 | 1.75 | 2.62 |
| I10 | 66.02 | 67.34 | 63.87 | 50.35 | 51.71 | 48.13 |
| I48 | 32.44 | 31.28 | 34.35 | 24.3 | 23.78 | 25.16 |
| S06 | 9.03 | 8.86 | 9.29 | 4.22 | 4.2 | 4.24 |
| F32-F33 | 17.08 | 19.82 | 12.63 | 5.6 | 7.06 | 3.23 |
| G45-G46 | 5.06 | 4.91 | 5.3 | 2.99 | 3.02 | 2.93 |
| I10-I15 | 66.53 | 67.86 | 64.36 | 50.87 | 52.23 | 48.66 |
| E10-E14 | 24.33 | 22.66 | 27.05 | 15.34 | 14.37 | 16.9 |
| I20-I25 | 24.84 | 21.65 | 30.03 | 21.07 | 17.73 | 26.5 |
| H90-H91 | 4.9 | 4.67 | 5.27 | 2.35 | 2.28 | 2.46 |

**TABLE V:** Prevalence of identified dementia risk factors in patients with dementia (ICD-10: F00, F01, F02, F03, G30) and matched controls, paired by age, sex, and year of diagnosis.

| ICD | Condition | Dementia |  |  | Without Dementia |  |  |
| --- | --- | --- | --- | --- | --- | --- | --- |
|  |  | All | Female | Male | All | Female | Male |
| A04 | Bacterial intestinal infections | 2.01 | 2.01 | 2.01 | 1.01 | 1.09 | 0.87 |
| A09 | Infectious gastroenteritis and colitis | 4.76 | 5.21 | 4.03 | 2.72 | 3.09 | 2.10 |
| B37 | Candidiasis | 1.60 | 1.47 | 1.82 | 0.78 | 0.76 | 0.82 |
| B99 | Infectious diseases | 4.69 | 4.08 | 5.67 | 1.76 | 1.59 | 2.03 |
| E03 | Hypothyroidism | 9.24 | 11.36 | 5.80 | 5.69 | 7.22 | 3.21 |
| E11 | Type 2 diabetes mellitus | 22.69 | 21.09 | 25.30 | 13.92 | 13.05 | 15.34 |
| E14 | Diabetes mellitus | 5.32 | 4.75 | 6.25 | 2.77 | 2.49 | 3.24 |
| E53 | Vitamin B deficiency | 4.04 | 3.85 | 4.34 | 0.96 | 0.94 | 1.00 |
| E55 | Vitamin D deficiency | 5.06 | 5.29 | 4.69 | 2.16 | 2.45 | 1.68 |
| E78 | Disorders of lipoprotein metabolism | 19.63 | 18.82 | 20.96 | 15.43 | 14.66 | 16.68 |
| E86 | Volume depletion | 20.35 | 21.23 | 18.93 | 5.62 | 6.23 | 4.64 |
| E87 | Fluid, electrolyte and acid-base disorders | 14.28 | 15.56 | 12.18 | 6.67 | 7.82 | 4.80 |
| F00 | Alzheimer's disease dementia | 22.61 | 22.67 | 22.51 | 0.00 | 0.00 | 0.00 |
| F01 | Vascular dementia | 21.41 | 20.62 | 22.68 | 0.00 | 0.00 | 0.00 |
| F02 | Dementia in other diseases | 3.45 | 2.45 | 5.07 | 0.00 | 0.00 | 0.00 |
| F03 | Dementia | 62.48 | 63.01 | 61.61 | 0.00 | 0.00 | 0.00 |
| F05 | Delirium | 9.03 | 7.81 | 11.02 | 1.07 | 0.91 | 1.34 |
| F06 | Organic mental disorders | 2.81 | 2.60 | 3.16 | 0.65 | 0.69 | 0.59 |
| F07 | Personality and behavioural disorders due to brain disease | 1.39 | 1.01 | 2.01 | 0.17 | 0.12 | 0.24 |
| F10 | Alcohol-related mental and behavioural disorders | 3.00 | 1.47 | 5.48 | 0.65 | 0.25 | 1.30 |
| F32 | Depressive episode | 14.33 | 16.65 | 10.56 | 4.82 | 6.07 | 2.78 |
| F33 | Recurrent depressive disorder | 5.22 | 6.15 | 3.73 | 1.33 | 1.71 | 0.71 |
| G20 | Parkinson disease | 8.98 | 6.76 | 12.59 | 2.08 | 1.75 | 2.62 |
| G21 | Secondary parkinsonism | 1.94 | 1.47 | 2.71 | 0.29 | 0.26 | 0.35 |
| G30 | Alzheimer disease | 35.28 | 35.70 | 34.61 | 0.00 | 0.00 | 0.00 |
| G31 | Other neurodegenerative disorders | 2.15 | 1.80 | 2.72 | 0.25 | 0.25 | 0.24 |
| G40 | Epilepsy | 7.32 | 6.77 | 8.21 | 1.71 | 1.61 | 1.88 |
| G62 | Polyneuropathy | 4.51 | 3.70 | 5.84 | 2.47 | 2.37 | 2.65 |
| G91 | Hydrocephalus | 1.02 | 0.73 | 1.49 | 0.17 | 0.14 | 0.22 |
| G93 | Other brain disorders | 3.42 | 3.24 | 3.73 | 0.92 | 0.93 | 0.90 |
| I10 | Essential hypertension | 66.02 | 67.34 | 63.87 | 50.35 | 51.71 | 48.13 |
| I48 | Atrial fibrillation and flutter | 32.44 | 31.28 | 34.35 | 24.30 | 23.78 | 25.16 |
| I50 | Heart failure | 22.74 | 22.45 | 23.21 | 16.58 | 16.74 | 16.32 |
| I67 | Cerebrovascular diseases | 10.10 | 9.40 | 11.24 | 3.81 | 3.83 | 3.77 |
| I69 | Sequelae of cerebrovascular disease | 6.58 | 5.53 | 8.29 | 2.18 | 2.06 | 2.40 |
| I70 | Atherosclerosis | 9.94 | 8.80 | 11.80 | 6.81 | 6.22 | 7.76 |
| I80 | Phlebitis and thrombophlebitis | 3.19 | 3.47 | 2.75 | 1.89 | 2.02 | 1.68 |
| J15 | Bacterial pneumonia | 7.89 | 6.06 | 10.86 | 3.63 | 3.02 | 4.63 |
| J18 | Pneumonia | 21.22 | 17.48 | 27.30 | 9.99 | 8.88 | 11.79 |
| J20 | Acute bronchitis | 3.73 | 3.53 | 4.04 | 2.12 | 2.24 | 1.92 |
| J22 | Acute lower respiratory infection | 2.30 | 2.05 | 2.71 | 1.08 | 1.08 | 1.08 |
| J40 | Bronchitis | 2.25 | 2.20 | 2.35 | 1.17 | 1.26 | 1.03 |
| J69 | Aspiration pneumonitis | 4.77 | 3.37 | 7.05 | 1.00 | 0.79 | 1.35 |
| K59 | Functional intestinal disorders | 7.05 | 7.04 | 7.06 | 3.63 | 3.89 | 3.22 |
| L89 | Pressure ulcer | 3.99 | 4.16 | 3.72 | 1.04 | 1.13 | 0.91 |
| M81 | Osteoporosis | 15.60 | 21.40 | 6.16 | 9.19 | 12.93 | 3.13 |
| N17 | Acute kidney failure | 9.96 | 9.17 | 11.24 | 5.90 | 5.58 | 6.40 |
| N18 | Chronic kidney disease | 25.94 | 25.51 | 26.63 | 15.94 | 15.64 | 16.42 |
| N19 | Kidney failure | 7.46 | 7.29 | 7.74 | 3.98 | 3.89 | 4.12 |
| N30 | Cystitis | 1.80 | 1.89 | 1.67 | 0.83 | 0.87 | 0.78 |
| N39 | Urinary system disorders | 33.39 | 37.70 | 26.38 | 14.03 | 16.71 | 9.67 |

**Fig. 1:**
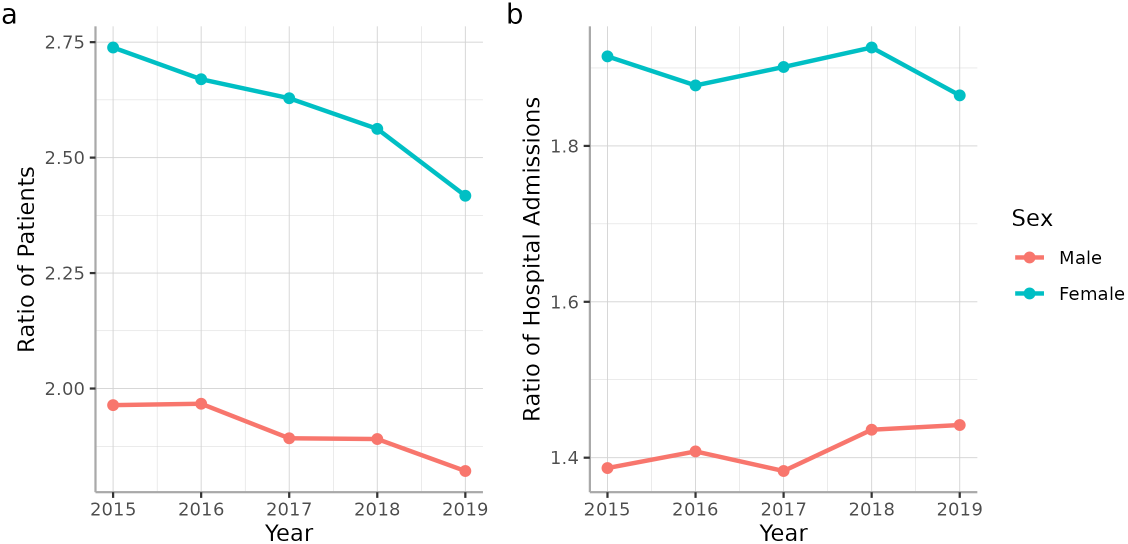
a) Prevalence of dementia and b) hospitalization admissions with dementia for females and males over time

**Fig. 2:**
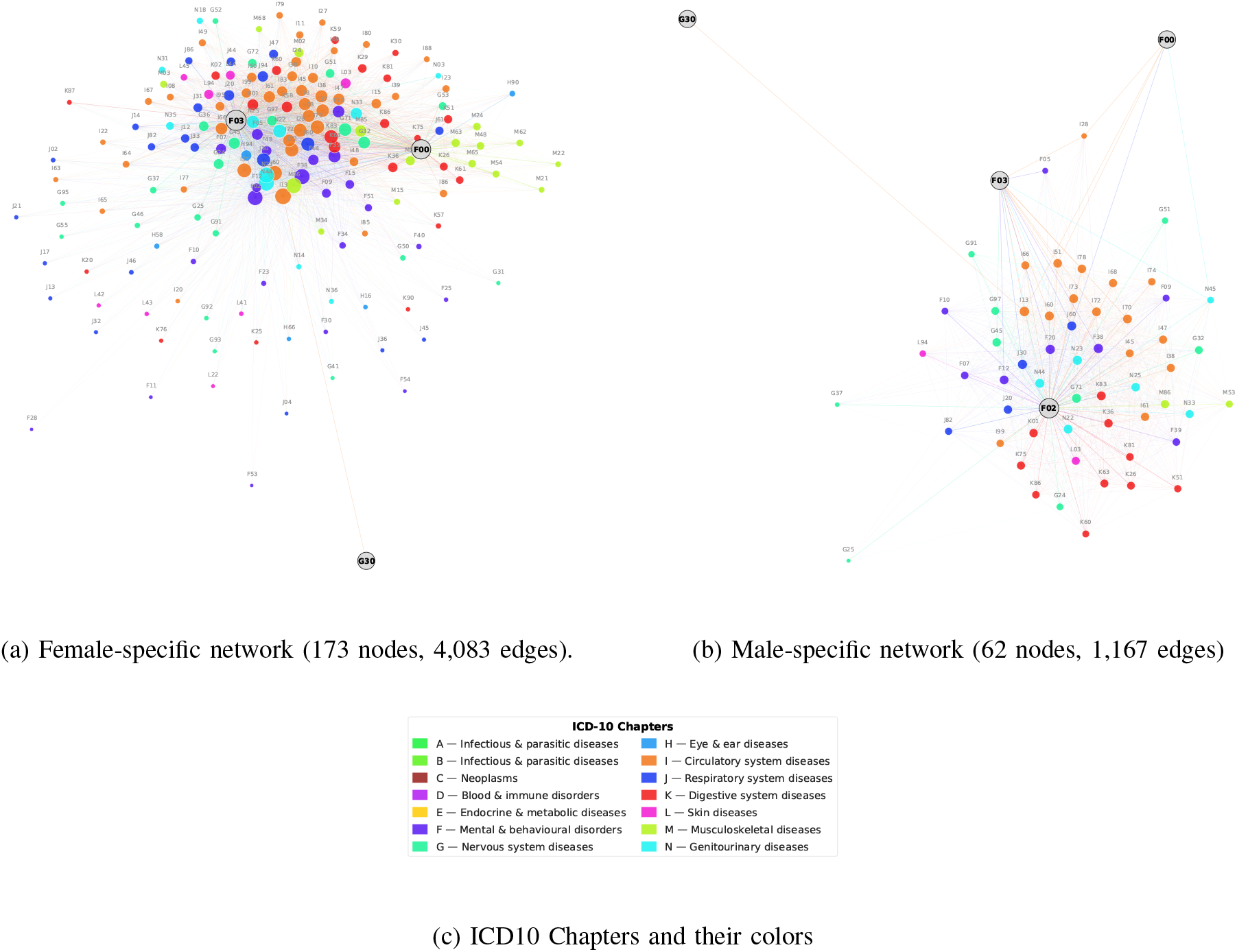
Sex-specific dementia comorbidity networks constructed from Austrian nationwide hospital claims data (2015–2019). Nodes represent ICD-10 diagnoses; edges indicate statistically significant co-occurrence with at least one dementia code. Node color reflects ICD-10 chapter as shown in the legend and node size is proportional to the degree.

### A. Dementia Comorbidities Through Comorbidity Networks

Having observed this sex-specificity, we compared the features of the dementia-specific co-occurrence networks, Figure 3. The male network exhibited higher density, resulting in higher centrality measures (degree, betweenness, K-core), even after controlling for differences in network size. The interpretation of this finding is that, on average, men with dementia tend to be afflicted by a higher number of comorbidities, which is aligned with the higher number of diagnoses and hospitalizations observed in Table IV. The vast majority of male disease co-occurrences were also present in females (86%, number of links=1,007), as shown by the most connected nodes, Figure 4.Hypertensive heart and chronic kidney disease (I13) was the most connected node in both genders. Cardiovascular diseases (Chapter I) and genitourinary diseases (Chapter N) were the most reported disorders in dementia patients. Interestingly, nonrheumatic pulmonary valve disorders (I37) occurred more frequently in women than in men, which is in agreement with epidemiological observation in the American population [51].

**Fig. 3:**
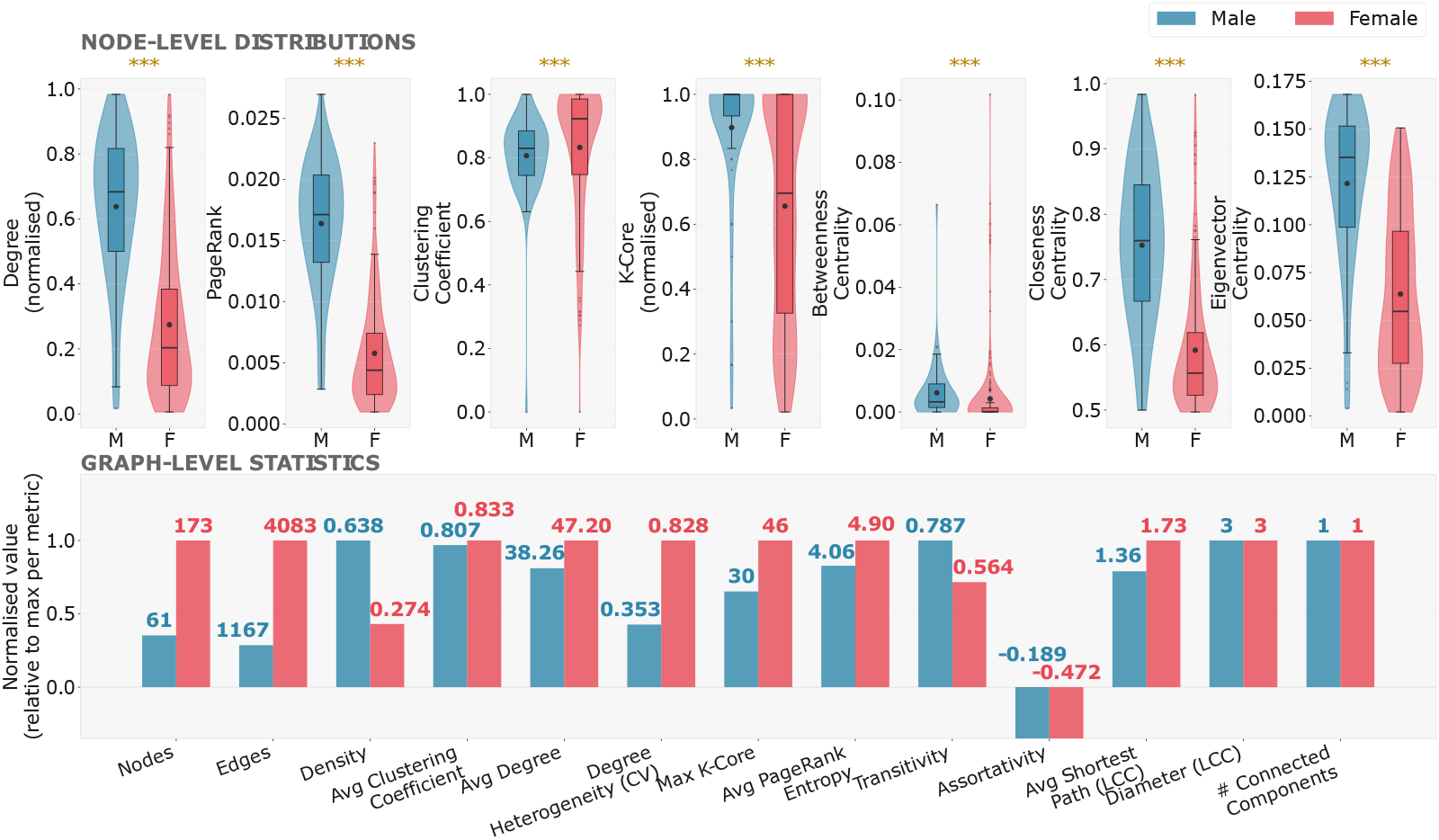
Kolmogorov–Smirnov (KS) test significance: ***p < 0.001, **p < 0.01, *p < 0.05, ns = not significant. Graph-level bars show values normalised to the larger of the two networks per metric; raw values are annotated above each bar.

**Fig. 4:**
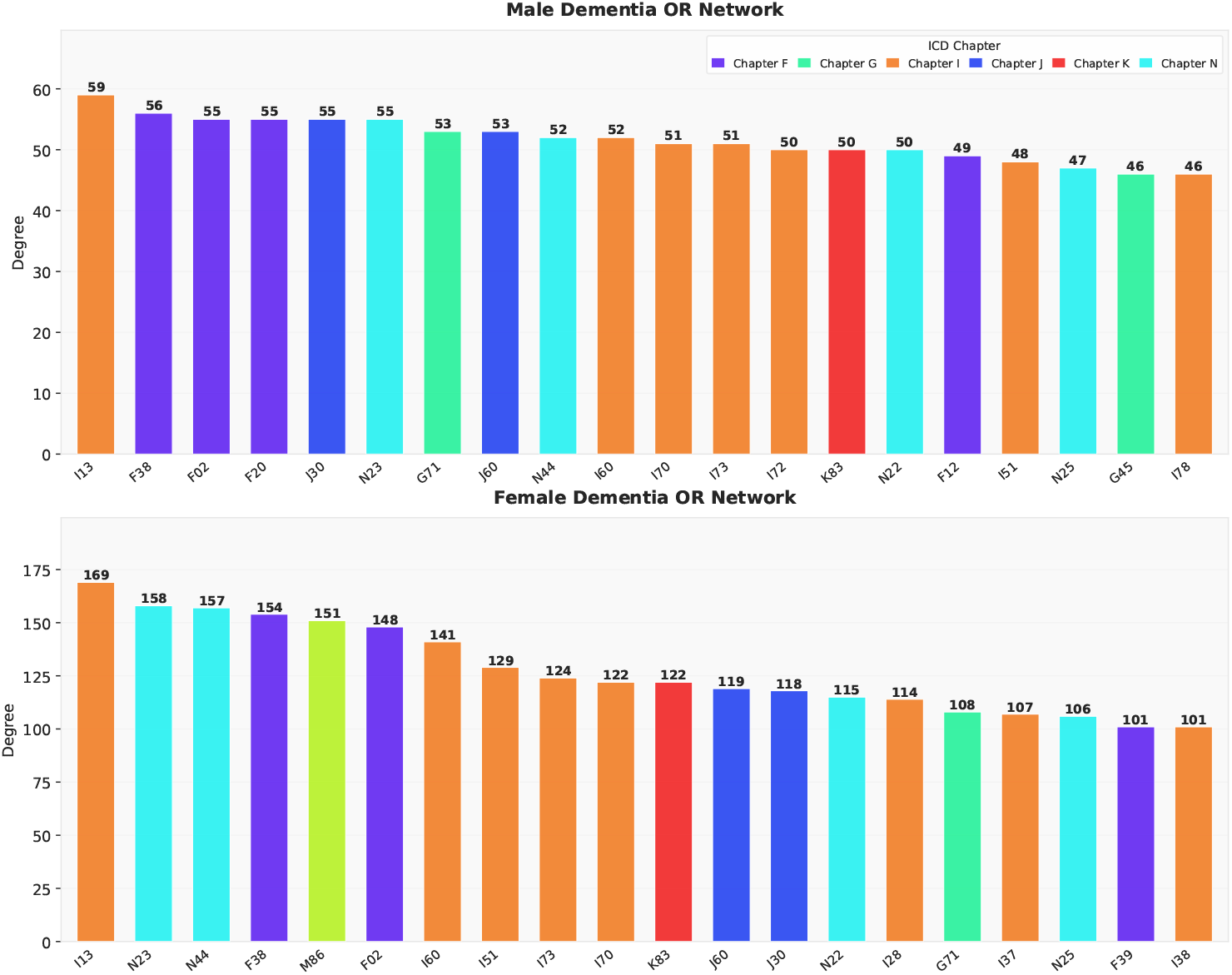
Top 20 most connected nodes (by degree) in the male (upper) and female (lower) dementia comorbidity networks. Bar colors indicate ICD-10 chapter. Degree values are annotated above each bar.

#### 1) Comorbidity Clusters of Dementia

In both sex-specific dementia-specific comorbidity networks the Louvain communities identified three major groups in men and four in women. The disease representation across these communities was quite heterogeneous, with only two community pairs sharing a statistically significant portion of diseases across genders (male community 2 with female community 3, FDR = 0.009; and male community 1 with female community 2, FDR = 0.01; hypergeometric test with Benjamini-Hochberg method for multiple comparisons), Table 1 and 2 in the SI.

#### 2) Dementia Risk Factors

While the exploration of the comorbidity patterns associated with dementia provides important information regarding the diseases co-occurring with dementia, it is equally important to study those diseases occurring before developing dementia. For this purpose, we collected diseases with a statistically significantly higher incidence in dementia patients (compared with matched controls) over the 5 years preceding dementia diagnosis, and the co-occurring diseases in the 2 years preceding dementia in both genders. With this approach, we obtained an overview of 51 diseases and their co-occurrences which could lead to dementia before diagnosis, as shown in Figure 5a. Interestingly, only atherosclerosis (I70) was a risk factor that also occurred in the dementia-specific comorbidity networks, highlighting the importance of this approach for tracing back possible risk factors in the past. Most risk factors were shared between genders, except for phlebitis and thrombophlebitis (I80), which were specific to women. The most co-occurring diseases before developing dementia were pneumonia (J18) and depression (F32). These associations have been confirmed from previous studies, reporting a 2-fold or greater increase in risk of dementia for people suffering from depression [52], and an estimated 70% increase in risk with pneumonia [53]. Unspecified dementia (F03) showed the highest number of disease risk factors (n = 39), and Alzheimer’s disease (G30) was the second (n = 24). Interestingly, endocrine, nutritional, and metabolic diseases (Chapter E) were the most representative class of risk factors for dementia, Figure 5b, with vitamin D deficiency (E55) being the most connected disease in this class. This result is not surprising and has already been reported in other, smaller cohorts [54]. Other neurological disorders (Chapter G) have resulted in a strong risk factor for dementia, including epilepsy (G40, n=4) and Parkinson’s disease (G20, n=3).

**Fig. 5:**
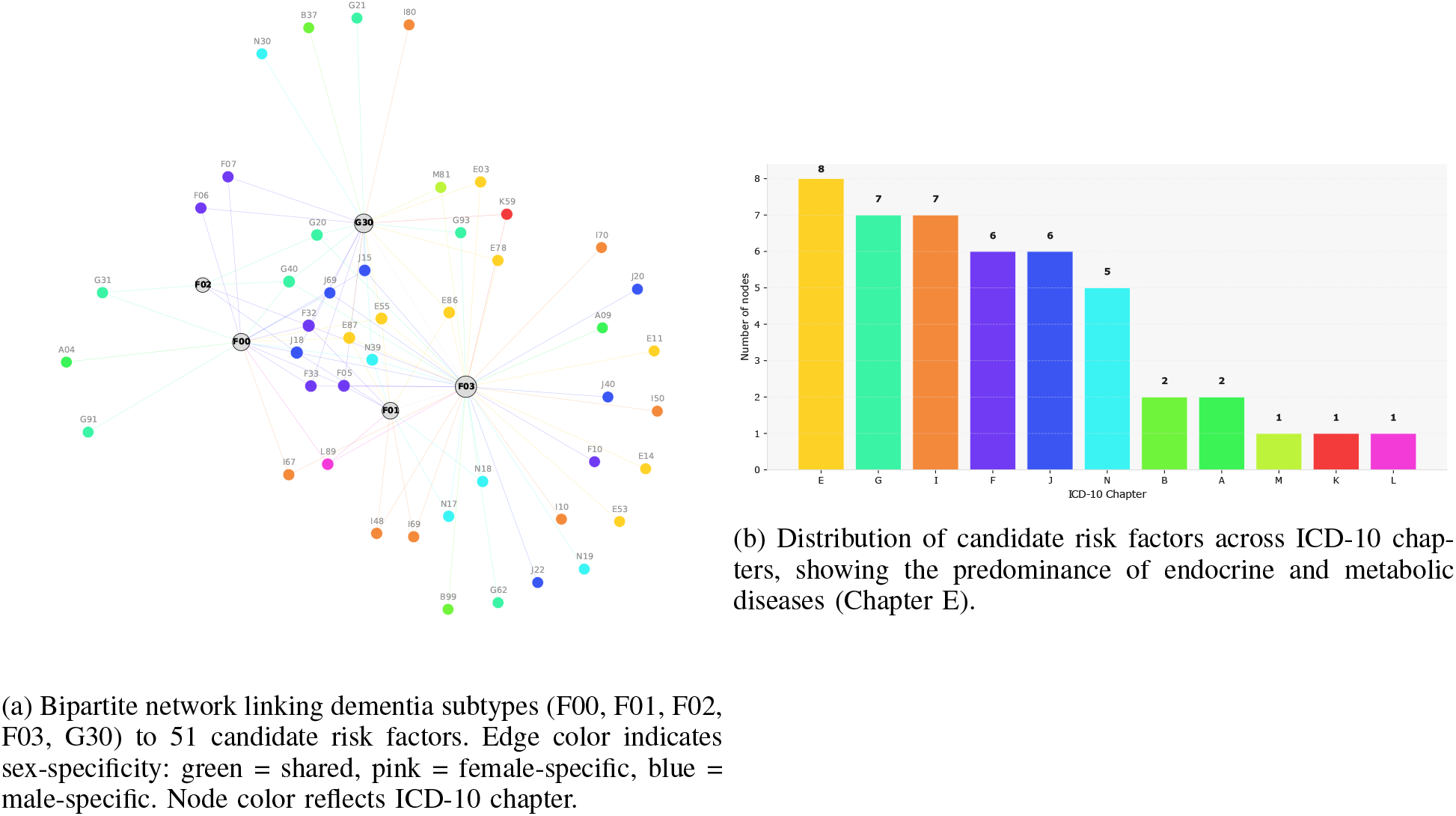
Dementia risk factors identified in the 5 years preceding diagnosis.

### 3) Assessing Genetic Comorbidity Signatures in Clinical Dementia Records

We constructed a disease–disease correlation matrix based on gene–disease associations extracted from the OpenTargets platform’s database. This matrix was subsequently harmonized with clinical data using ICD-10 disease codes. The harmonization was performed by mapping disease names and comparing embedding similarities derived from the PubMedBert model [48]. Focusing on dementia-related conditions, grouped under umbrella term Dementia (F00), and Alzheimer’s Disease (G30), we extracted the top 50 genetically associated diseases from the constructed matrix. We calculated the Odds Ratio (OR) and statistical significance of the genetic overlap through a hypergeometric test with adjustment for multiple comparisons.

First, we assessed whether diseases co-occurring with dementia were also genetically related. We observed that diseases co-occurring with dementia shared in both genders had the highest genetic odds ratio (median OR = 2.8; Figure 6a), with the strongest molecular association retrieved with unspecified renal colic (N23; OR=11.9). Interestingly, also female-specific edges had a strong molecular similarity, such as schizoaffective disorders (F25; OR = 13.9). Almost all comorbidity communities identified in both genders showed an average genetic OR approximately equal to 2, without any prominent differences among them, Figure 6b. We then moved to evaluate the amount of shared genetic component between the dementia risk factors and dementia itself (both general dementia and Alzheimer’s disease), and we corroborated their association by collecting the number of PubMed articles co-mentioning them. Figure 7. In Alzheimer’s disease, Parkinson’s disease was observed to share a high molecular similarity (FDR<1e300, Fisher’s exact test with Benjamini-Hochberg correction), as shown also by the high number of PubMed articles co-mentioning those two conditions (n=9,999).

**Fig. 6:**
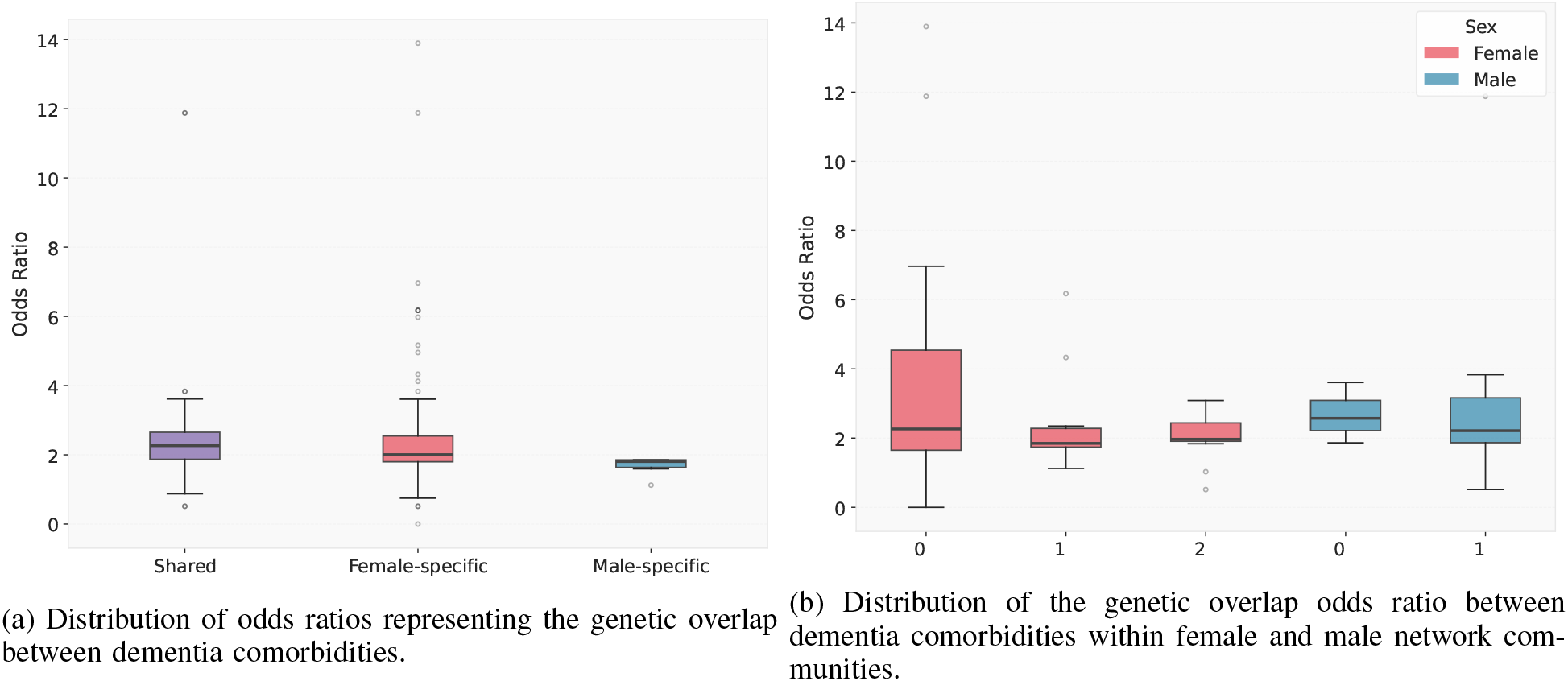
Genetic overlap between dementia comorbidities extracted from the sex-specific dementia comorbidity networks

**Fig. 7:**
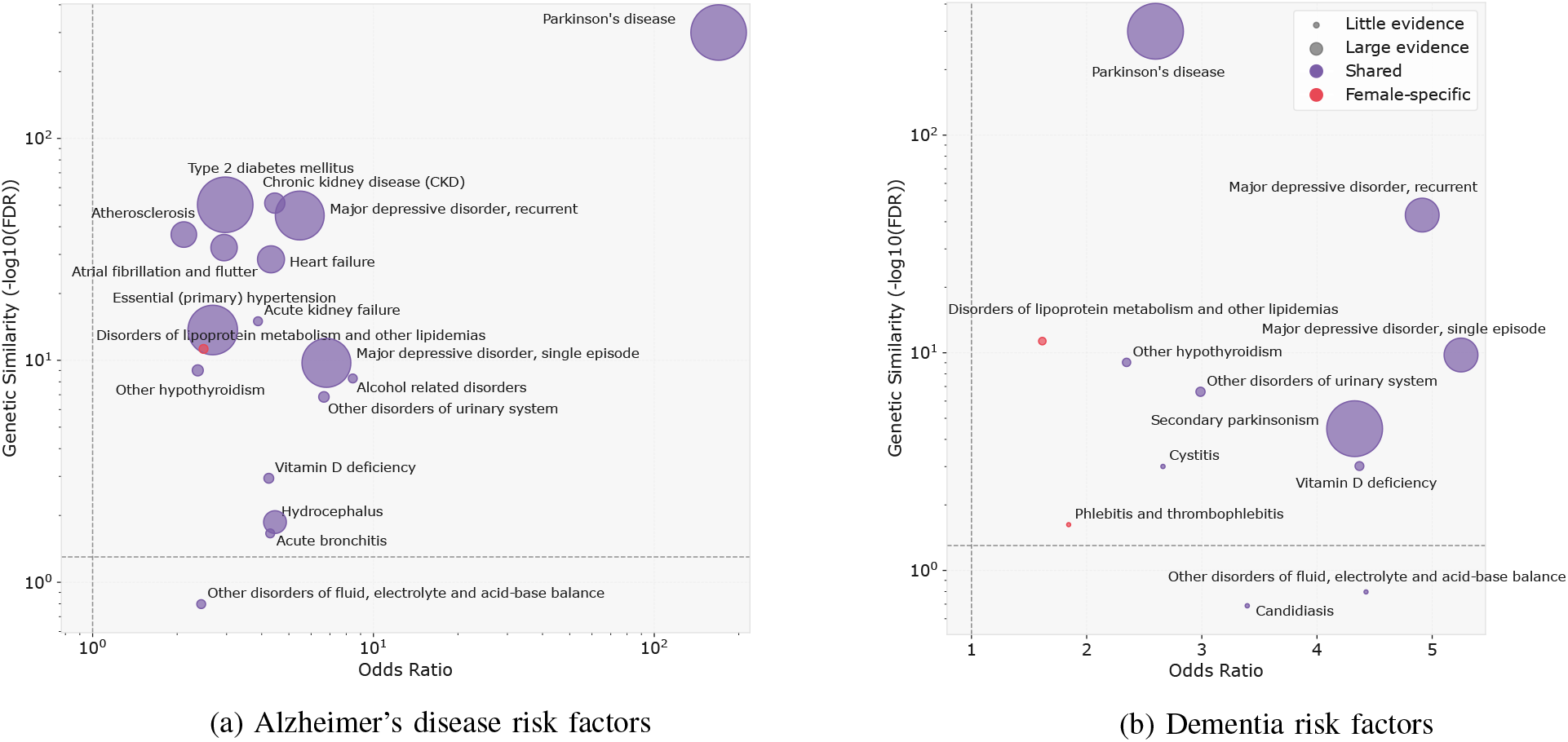
Clinical and molecular similarity of disease risk factors with Alzheimer’s disease and dementia

On the other hand, the association with Vitamin D deficiency was less reported in the literature (number of co-mentions = 196), but it showed a strong clinical association (OR=4.4) and molecular similarity (FDR = 9e-5). Across all dementia risk factors, Parkinson’s disease remains one of the most prominent associations (clinical OR = 169, genetic FDR < 1e-300, number of co-mentions = 9,999), but also some lifestyle-related risk factors, such as alcohol-related disorders (clinical OR = 8, genetic FDR = 5e-09, number of co-mentions = 79), emerge as important preventable interventions. Finally, we ranked potential disease risk factors based on their molecular similarity with overall dementia and Alzheimer’s disease, confirming Parkinson’s disease as the most relevant one from a molecular standpoint, and highlighting the contribution of diseases from other categories, such as Type 2 diabetes mellitus, and atherosclerosis, Figure 9.

**Fig. 8:**
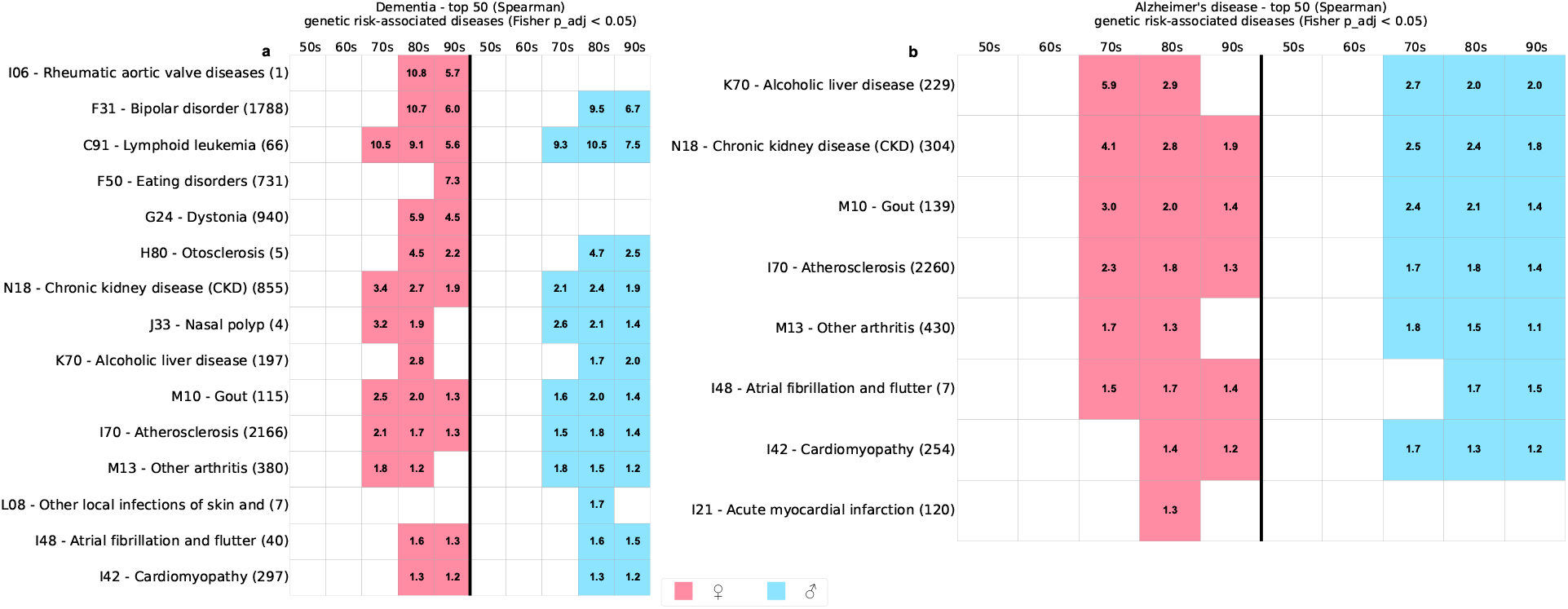
Genetic comorbidity signatures validated in Austrian hospital claims data. Panels show a subset of the top 100 genetically associated diseases (from Open Targets, after mapping to ICD-10) that were also observed to co-occur in clinical records with (a) Dementia (F00) and (b) Alzheimer’s disease (G30). The grid indicates in which age decades (50s–90s) and sex the co-occurrence was detected (red - females, blue - males). Numbers in parentheses next to each ICD-10 label and disease name denote the number of PubMed articles mentioning the disease together with dementia/Alzheimer’s disease, used as supporting literature evidence.

**Fig. 9:**
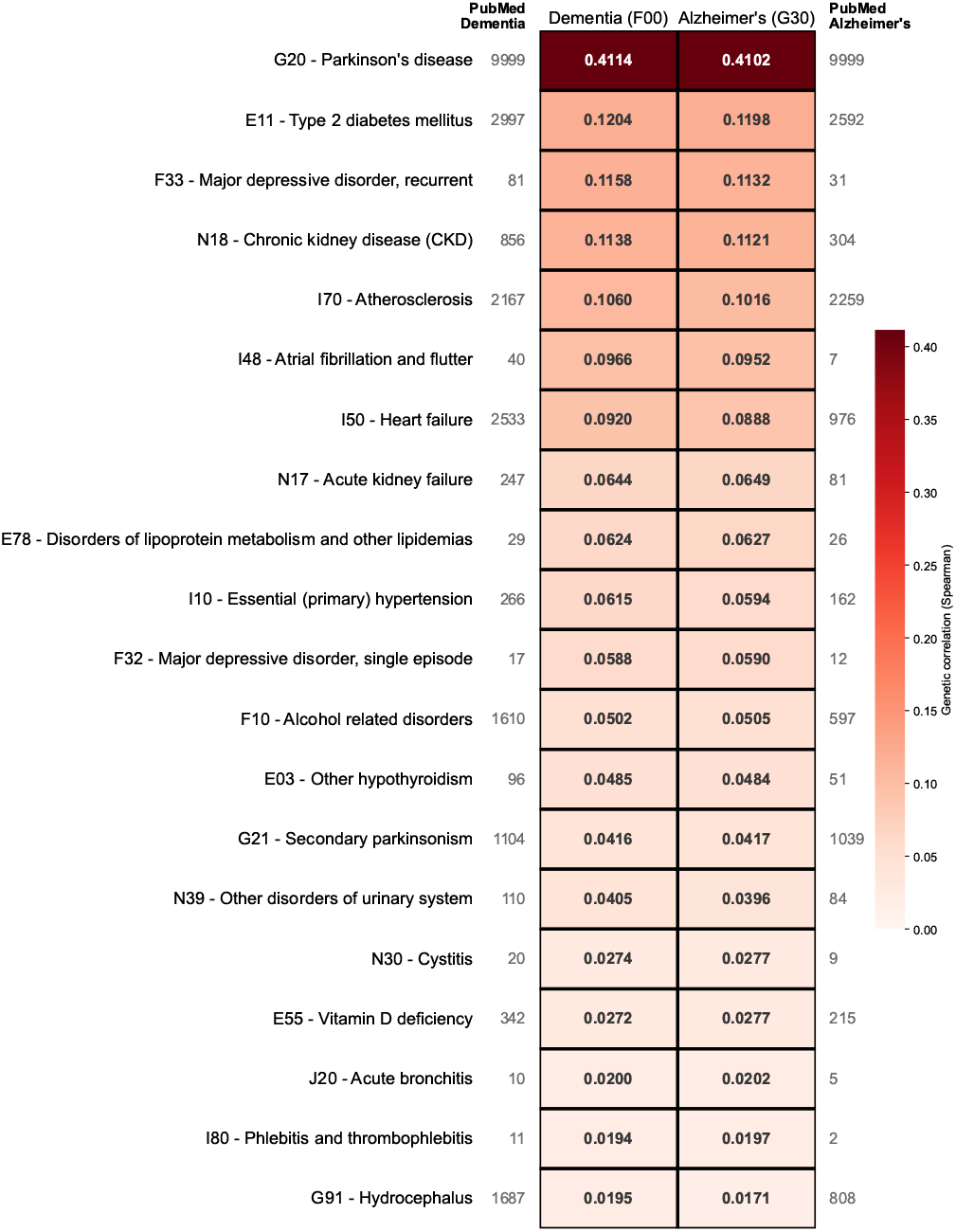
Genetic correlation between selected dementia risk factors and two dementia conditions: Dementia (F00) and Alzheimer’s disease (G30). The heatmap displays Spearman correlation coefficients derived from shared gene associations in the OpenTargets database. Only diseases with a maximum absolute correlation ≥ 0.01 are shown (20 out of 46 selected risk factors). Cells with a black border indicate statistically significant gene overlap as determined by Fisher’s exact test (Benjamini–Hochberg adjusted p < 0.05). The number of co-occurring publications in PubMed is shown on the left for Dementia and on the right for Alzheimer’s disease, providing an independent measure of the known association between each risk factor and the respective dementia condition. The color intensity reflects the strength of the genetic correlation, with darker shades indicating stronger shared genetic risk architecture.

## V. Discussion

Dementia represents a major and growing global health challenge that requires timely intervention, with prevention increasingly recognized as a key priority. In this study, we implemented a quantitative, data-driven approach to characterize disease comorbidities occurring both at and prior to dementia diagnosis. This enabled the identification of sex-specific as well as shared comorbidity patterns that might inform preventive strategies. We found that the majority of disease co-occurrences at and preceding dementia diagnosis are shared between sexes, recapitulating established associations with conditions such as diabetes, depression, dyslipidemia, and excessive alcohol use. Beyond confirming known risk factors, we identified a total of 51 potential risk factors, with a predominance of endocrine and metabolic disorders, as well as neurological and cardiovascular conditions. For instance, our findings suggest that vitamin D deficiency may represent an additional contributing factor in dementia development.

A prominent finding is the marked sex asymmetry in comorbidity patterns. The female network was substantially larger than the male network (173 vs. 62 nodes), yet 86% of male co-occurrences were also present in the female network, suggesting a broadly shared biological substrate. Several factors likely contribute to this asymmetry. Women have a longer life expectancy in Austria, resulting in an older female dementia population (mean age 87.07 vs. 84.38 years) with more time to accumulate comorbidities [50]. Hormonal factors, particularly post-menopausal estrogen decline, may further shape the female comorbidity profile. Despite their smaller network, male patients showed higher network density and centrality, consistent with a more tightly interconnected network and a greater comorbidity burden, as reflected in more hospital stays and diagnoses, Table IV.

Importantly, we integrated clinical data with measures of molecular similarity between disease pairs. This enabled distinguishing associations driven by shared biological mechanisms from those potentially arising through environmental or non-molecular pathways. For instance, our findings show a strong molecular association between dementia and Parkinson’s disease. They also highlight new potential risk factors, such as vitamin D deficiency, significantly molecularly similar to Alzheimer’s disease, despite the limited existing literature.

## VI. Strength and limitations

Despite its strengths, this study has several limitations. First, it captures only inpatient care and lacks information on outpatient visits, medication use, and lifestyle factors. Moreover, the analysis is restricted to hospital diagnoses coded using ICD-10, introducing a bias toward more severe disease and potentially underrepresenting early or milder stages of dementia and related comorbidities managed outside hospital settings. Second, as the database was primarily designed for billing purposes, diagnoses not affecting reimbursement may be incompletely recorded. Third, coding errors and limited clinical granularity may impact subtype-specific analyses. Nevertheless, the number of dementia cases identified in our study is comparable to previous reports based on national prescription data in Austria, supporting the validity of the overall dementia burden captured in this dataset [17].

## Data Availability

The raw and processed patient data are not available due to privacy laws. The dataset is safeguarded by the Austrian Federal Ministry of Health and made accessible to research institutions under strict data protection regulations. To gain access to this data, researchers have to find individual arrangements with the Austrian Federal Ministry of Health.

## VII. Ethics

We used a securely managed medical claims research database overseen by the Federal Ministry of Health. The database contains no personal identifiers and is accessible only to authorized partners under strict data protection policies. Our use of the data was conducted in accordance with established agreements, and team members, who worked with the data, signed confidentiality commitments in compliance with relevant data protection regulations.

## VIII. Data Availability

Disease-disease network extracted from https://platform.opentargets.org/ disease gene associations is publicly available.

## IX. Code Availability

Custom code for the analysis is available per request from the authors. The code used for downloading and preparing Open Targets disease-disease matrix, harmonize it with hospital data and creating visualization is publicly available at https://github.com/vladimirkovacevic/CSH/.

## X. Acknowledgment

This work was supported by the Ministry of Science, Technological Development and Innovation of the Republic of Serbia (No. 337-00-216/2023-05/268) and by OeAD-GmbH on behalf of the Austrian Federal Ministry of Education, Science and Research (No. RS 23/2024), through the Serbia–Austria WTZ bilateral cooperation program 2024–2026.

---

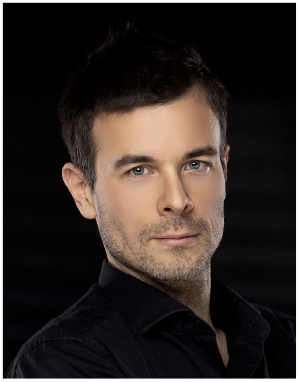

**Vladimir Kovačević** received the Ph.D. degree in computer science and has more than 18 years of experience in scientific software engineering and algorithm development, with over a decade of work in bioinformatics, genomics, and artificial intelligence. His research focuses on machine learning for biological data, multiomics integration, spatial transcriptomics, immuno-oncology, application of large language models to protein sequences and construction of large-scale cloud-based bioinformatics systems.

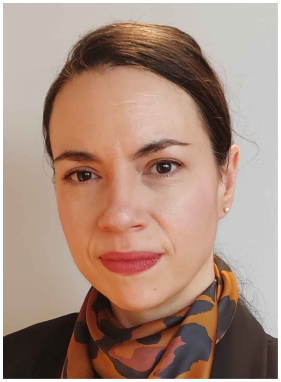

**Bojana Bašaragin** received her Bachelor’s degree in General Linguistics in 2006 and her Doctor of Philosophy (Ph.D.) in Computational Linguistics from the Faculty of Philology, University of Belgrade in 2017. She is currently a Senior Researcher at the Institute for Artificial Intelligence Research and Development of Serbia. Her work focuses on natural language processing (NLP) applied to Serbian language data, particularly the creation and maintenance of linguistic resources and tools using large language models (LLMs). She has been involved in both academic and industrial projects, ranging from healthcare-related NLP to the development of pipelines for the improvement of customer service in companies. Dr. Bašaragin is a founding member of the Serbia-based Language Resources and Technologies Society.

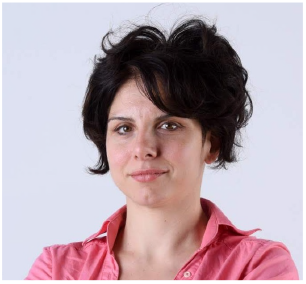

**Jovana Kovačević** earned her BSc (2007) and PhD (2015) in Computer Science from the Faculty of Mathematics, University of Belgrade, with doctoral research at the intersection of bioinformatics, data mining, and machine learning, focusing on computational analysis of biological data and protein structure and function. She is currently an Associate Professor at the same faculty. In addition to her academic role, she has worked as a Research Associate at the Institute for Artificial Intelligence of Serbia within the AI in Healthcare and Life Sciences group, as well as a consultant for the Centre for the Fourth Industrial Revolution, and has also taught at the Mathematical High School in Belgrade. Her research interests include bioinformatics, machine learning, and text and data mining, with emphasis on intrinsically disordered proteins, protein function prediction, and AI applications in life sciences and healthcare.

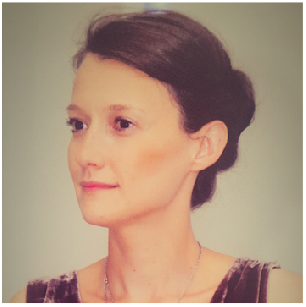

**Anđelka Zečević** is a researcher at the Mathematical Institute of Serbian Academy of Sciences and Arts. She is completing Ph.D. studies at the Faculty of Mathematics, University of Belgrade, at the Department for Computer Science and Informatics in the domain of Natural Language Processing. Her research focuses on the language of medicine and the development of algorithms that enhance the understanding of complex medical knowledge. She also works on data-driven algorithms aimed at advancing precision medicine and improving patient safety.

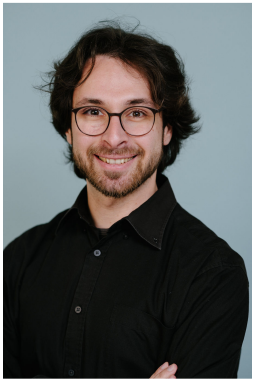

**Salvo Danilo Lombardo**, MD, is a Postdoctoral Fellow at the Channing Division of Network Medicine, Brigham and Women’s Hospital, Harvard Medical School. He earned his medical degree with honors from the University of Catania in 2019 and completed his PhD at the Medical University of Vienna and at CeMM – the Research Center for Molecular Medicine of the Austrian Academy of Sciences in 2025. During his PhD, Salvo was also a Predoctoral Fellow at the Max Perutz Labs (Vienna BioCenter) and at the Ludwig Boltzmann Institute for Network Medicine at the University of Vienna. Since 2023, he has been a Lindau Nobel Laureate Meetings Fellow. His research focuses on applying network science and network medicine approaches to environmental-genetic interactions to better understand disease origins.

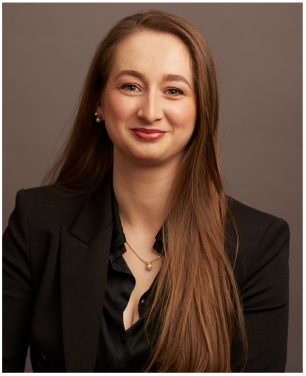

**Elma (Hot) Dervic** is a postdoctoral fellow at the Complexity Science Hub and the Medical University of Vienna. She completed her PhD at CSH and the Medical University of Vienna in January 2023. Elma holds a bachelor’s and a master’s degree in electrical engineering from the University of Montenegro, which she earned in 2015 and 2017, respectively. Before joining CSH, Elma was a junior researcher at the First Center of Excellence (BIO-ICT) at the University of Montenegro. Elma is a co-founder of Bee- And.me, a venture-funded IoT startup that uses technology to help beekeepers and bees through data science. She is known as a 3-time “share positive impact” TEDx speaker. Elma’s research interests include machine learning, network science, and everything impactful related to data science.

